# Nationwide Study on Clinical Outcomes of Endovascular Interventions for Cerebral Venous Thrombosis in Japan

**DOI:** 10.1101/2024.06.21.24309330

**Authors:** Atsushi Senda, Hiroshi Suginaka, Koji Morishita, Kiyohide Fushimi

## Abstract

**Background:** Cerebral venous thrombosis (CVT) is a rare but devastating disease, with some patients experiencing disease deterioration despite treatment. Endovascular treatment is an anticipated option, but its clinical relevance is yet to be determined. This observational study aimed to assess the clinical effects and identify patient populations that may benefit from treatment.

**Methods:** Patient data from April 2014 to March 2022 were extracted from a nationwide Japanese database. The primary outcome was in-hospital mortality, while secondary outcomes included modified Rankin Score (≥ 3) and posthospitalization complications. Severity adjustments were performed using a generalized linear mixed model and propensity score matching.

**Results:** The analysis included 2901 patients; 240 patients in the endovascular treatment group were matched with 240 patients in the standard treatment group. After adjusting for background factors, endovascular treatment did not improve in-hospital mortality (adjusted odds ratio 1.45 [95% CI: 0.74–2.16]) or the modified Rankin Score (adjusted odds ratio 0.89 [95% CI: 0.56–1.23]). No subpopulations that could benefit from endovascular treatment were identified. However, posthospitalization intracranial complications did not increase with endovascular treatment (0.8% vs. 1.2% in the standard treatment group).

**Conclusions:** Endovascular treatment did not show any clinical benefit in patients with CVT. These findings are crucial for guiding clinical decisions and suggest that further evidence is warranted.

## Introduction

Cerebral venous thrombosis (CVT), an uncommon cause of stroke, accounts for 0.5–3% of cerebrovascular diseases.^1–3^ It is a critical condition, with death or dependence occurring in 10–15% of patients, even after receiving intensive medical treatment.^2^

Anticoagulation is the cornerstone therapy for CVT,^4,5^ yet the condition of some patients deteriorates despite this treatment.^6–8^ Endovascular treatments, including mechanical thrombectomy, thromboaspiration, or balloon venoplasty with or without intrasinus thrombolysis, have been considered promising treatment options, supported by the findings of several case reports and systematic reviews.^9^ Nonetheless, the importance of endovascular treatments in CVT has dampened in recent years following the inability to demonstrate their efficacy in a randomized controlled trial (TO-ACT).^10^ However, abjuring endovascular treatments may be premature considering the low sensitivity of this trial conjointly with the feasibility and safety of this treatment option.^11^ Identifying the subpopulation who may benefit from this treatment is yet to be accomplished,^6^ and consequently, well-grounded indication criteria for endovascular treatments are not explicitly advocated in the current guidelines.^2,7,12^

Here, we conducted a nationwide retrospective observational study to reevaluate the treatment effect of endovascular therapy in patients with CVT with high sensitivity and identify the patient population that may benefit from these treatments.

## Methods

### Study design

In this study, we assessed the efficacy of endovascular treatments, including thrombectomy, angioplasty, fibrinolytic therapy, and stent placement, in patients with CVT using data from the Japanese Diagnosis Procedure Combination (DPC) database. In-hospital mortality was evaluated as the primary outcome. The DPC database, a nationwide case-mix patient classification system, lists over 1700 acute care hospitals, including all academic hospitals. The database not only contains assorted patient information, including age, sex, body weight, and underlying disease information, but also includes information on all procedures performed and routinely administered drugs.^13^ The diagnoses compiled in this database are in accordance with the International Classification of Diseases, Tenth Edition (ICD-10).^14^

This study adhered to the ethical standards of the Declaration of Helsinki and was approved by the Institutional Review Board of Tokyo Medical and Dental University (Approval No. 788, dated April 2020). As the study was retrospectively designed and deidentified data were used, the requirement to obtain informed consent was waived by the board.

### Study population

Inclusion criteria for the study included patients who were diagnosed with CVT (ICD-10 code: G08 “Intracranial and intraspinal phlebitis and thrombophlebitis”) and subsequently admitted to an intensive care unit during the designated study timeframe (April 2014–March 2022). Exclusion criteria included patients (i) with missing values for any of the variables utilized in the analysis, (ii) aged <16 years, and (iii) discharged within 2 d of admission. The latter criterion was used to address immortal time bias, as patient severity adjustment was executed according to the treatment intensity rendered during this period.

### Data collection

To appraise patient severity, the following variables were collected: concurrent diagnoses on admission, postadmission complications including cerebral infarction and intracranial hemorrhage, age, sex, Sequential Organ Failure Assessment (SOFA) score, and the state of consciousness upon admission. The following variables related to patient outcomes were also assessed: modified Rankin Scale (mRS), length of hospital stay, patient discharge status, and postadmission complications. Medications administered within 2 d postadmission including warfarin and other anticoagulants, antiepileptics, and antihypertensive drugs, were evaluated to gauge treatment intensity. Data on the year of admission and hospital identification number were also aggregated.

### Outcomes

The endovascular group included patients who underwent endovascular treatment within 2 d of admission. The primary outcome was in-hospital mortality, while the mRS and posthospitalization complications (intercranial hemorrhage and cerebral infarction) were assessed as secondary outcomes.

### Statistical analysis

To adjust for severity among patients with CVT who received endovascular treatment and those who did not, a prediction model for predicting patient prognosis was created. The model was constructed using a GLMM. The following explanatory variables were selected for the fixed effects of the model based on the findings of previous studies:^6,9^ age, sex, consciousness, cerebral infarction, intracranial hemorrhage, cerebral infarction, acute renal failure, heart failure, and use of drugs including warfarin, other anticoagulants, antiepileptics, and antihypertensive drugs. Hospital identification numbers were used for the random effects of the model.

The prediction model was constructed using 80% of a randomly selected cohort, while its accuracy was assessed on the remaining cohort using the bootstrap method (N = 1000). The accuracy was quantified by calculating the area under the receiver operating characteristic (AUROC) curve.

Sensitivity analysis was conducted using propensity score matching under the following conditions: 1:1 nearest neighbor matching without replacement, and a caliper width of 0.2 times the standard deviation of the logit-transformed propensity score. Comparisons between the matched groups were carried out using the Chi-square test.

For the main analysis, an antecedent generalized linear mixed model (GLMM) and propensity score matching were used. To identify the population of patients who may benefit from endovascular treatment, treatment effects were categorized according to the following three aspects: (1) patients stratified by disease severity, (2) patients stratified by the year of hospitalization, and (3) patients restricted to those aged <50 years. For the first analysis, patient severities were stratified using the prediction model, while treatment effects were estimated from the propensity score matching. For the latter two analyses, treatment effects were evaluated using the GLMM.

The R software, version 4.4.0 (R Foundation for Statistical Computing, Vienna, Austria) was used for performing all statistical analyses.

## Results

Figure 1 shows the patient selection process used in this study. During the study period, 7241 patients were diagnosed with CVT. A total of 3544 patients met the inclusion criteria, and the analysis was conducted in 2901 patients. Patients were excluded (n = 643) due to the following reasons: missing values (n = 443), aged <16 years (n = 83), pregnancy (n = 48), and discharge within 2 d of admission (n = 69). The baseline patient characteristics are presented in Table 1. The detailed treatments conducted in the endovascular treatment group were: thrombectomy, 161; angioplasty, 36; fibrinolytic therapy, 24; thrombectomy and angioplasty, 13; thrombectomy and fibrinolytic therapy, 3; thrombectomy and stent placement, 1; thrombectomy, angioplasty, and fibrinolytic therapy, 1; angioplasty and fibrinolytic therapy. 1. In the endovascular treatment group, the state of consciousness was poor, intracranial hemorrhage was frequently observed, and antiepileptic and antihypertensive drugs were often administered.

**Table 1.**
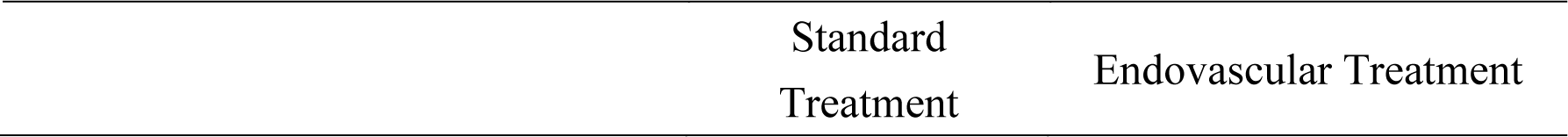
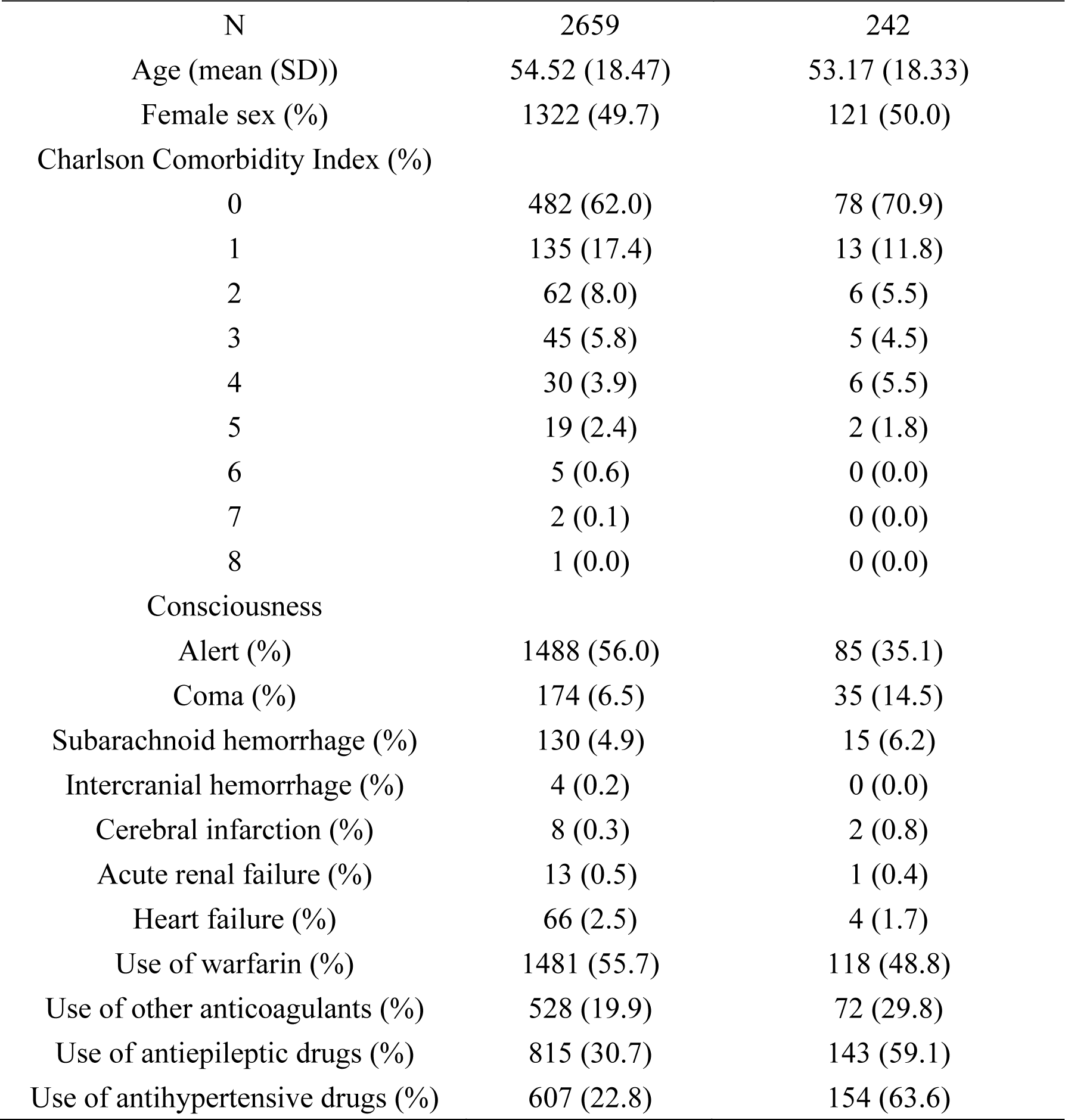
Patient characteristics.

|  | Standard<br>Treatment | Endovascular Treatment |
| --- | --- | --- |

| N | 2659 | 242 |
| --- | --- | --- |
| Age (mean (SD)) | 54.52 (18.47) | 53.17 (18.33) |
| Female sex (%) | 1322 (49.7) | 121 (50.0) |
| Charlson Comorbidity Index (%) |  |  |
| 0 | 482 (62.0) | 78 (70.9) |
| 1 | 135 (17.4) | 13 (11.8) |
| 2 | 62 (8.0) | 6 (5.5) |
| 3 | 45 (5.8) | 5 (4.5) |
| 4 | 30 (3.9) | 6 (5.5) |
| 5 | 19 (2.4) | 2 (1.8) |
| 6 | 5 (0.6) | 0 (0.0) |
| 7 | 2 (0.1) | 0 (0.0) |
| 8 | 1 (0.0) | 0 (0.0) |
| Consciousness |  |  |
| Alert (%) | 1488 (56.0) | 85 (35.1) |
| Coma (%) | 174 (6.5) | 35 (14.5) |
| Subarachnoid hemorrhage (%) | 130 (4.9) | 15 (6.2) |
| Intercranial hemorrhage (%) | 4 (0.2) | 0 (0.0) |
| Cerebral infarction (%) | 8 (0.3) | 2 (0.8) |
| Acute renal failure (%) | 13 (0.5) | 1 (0.4) |
| Heart failure (%) | 66 (2.5) | 4 (1.7) |
| Use of warfarin (%) | 1481 (55.7) | 118 (48.8) |
| Use of other anticoagulants (%) | 528 (19.9) | 72 (29.8) |
| Use of antiepileptic drugs (%) | 815 (30.7) | 143 (59.1) |
| Use of antihypertensive drugs (%) | 607 (22.8) | 154 (63.6) |

**Table 2.** Patient characteristics after propensity score matching.

|  | Standard<br>Treatment | Endovascular<br>Treatment | SMD |
| --- | --- | --- | --- |
| N | 240 | 240 |  |
| Age (mean (SD)) | 53.88 (17.57) | 53.27 (18.37) | 0.034 |
| Female sex (%) | 123 (51.2) | 120 (50.0) | 0.025 |
| Charlson Comorbidity Index (%) |  |  | 0.187 |
| 0 | 76 (31.7) | 71 (29.6) |  |
| 1 | 73 (30.4) | 80 (33.3) |  |
| 2 | 51 (21.2) | 53 (22.1) |  |
| 3 | 24 (10.0) | 26 (10.8) |  |
| 4 | 10 (4.2) | 7 (2.9) |  |
| 5 | 3 (1.2) | 3 (1.2) |  |
| 6 | 3 (1.2) | 0 (0.0) |  |
| Consciousness |  |  |  |
| Alert (%) | 84 (35.0) | 85 (35.4) | 0.009 |
| Coma (%) | 33 (13.8) | 35 (14.6) | 0.024 |
| Subarachnoid hemorrhage (%) | 24 (10.0) | 15 (6.2) | 0.138 |
| Intercranial hemorrhage (%) | 69 (28.7) | 71 (29.6) | 0.018 |
| Cerebral infarction (%) | 86 (35.8) | 78 (32.5) | 0.07 |
| Acute renal failure (%) | 1 (0.4) | 1 (0.4) | <0.001 |
| Heart failure (%) | 7 (2.9) | 4 (1.7) | 0.084 |
| Use of warfarin (%) | 122 (50.8) | 118 (49.2) | 0.033 |
| Use of other anticoagulants (%) | 72 (30.0) | 70 (29.2) | 0.018 |
| Use of antiepileptic drugs (%) | 136 (56.7) | 141 (58.8) | 0.042 |
| Use of antihypertensive drugs (%) | 153 (63.7) | 152 (63.3) | 0.009 |

Abbreviations: SMD, standardized mean difference, SD, standard deviation

**Figure 1:**
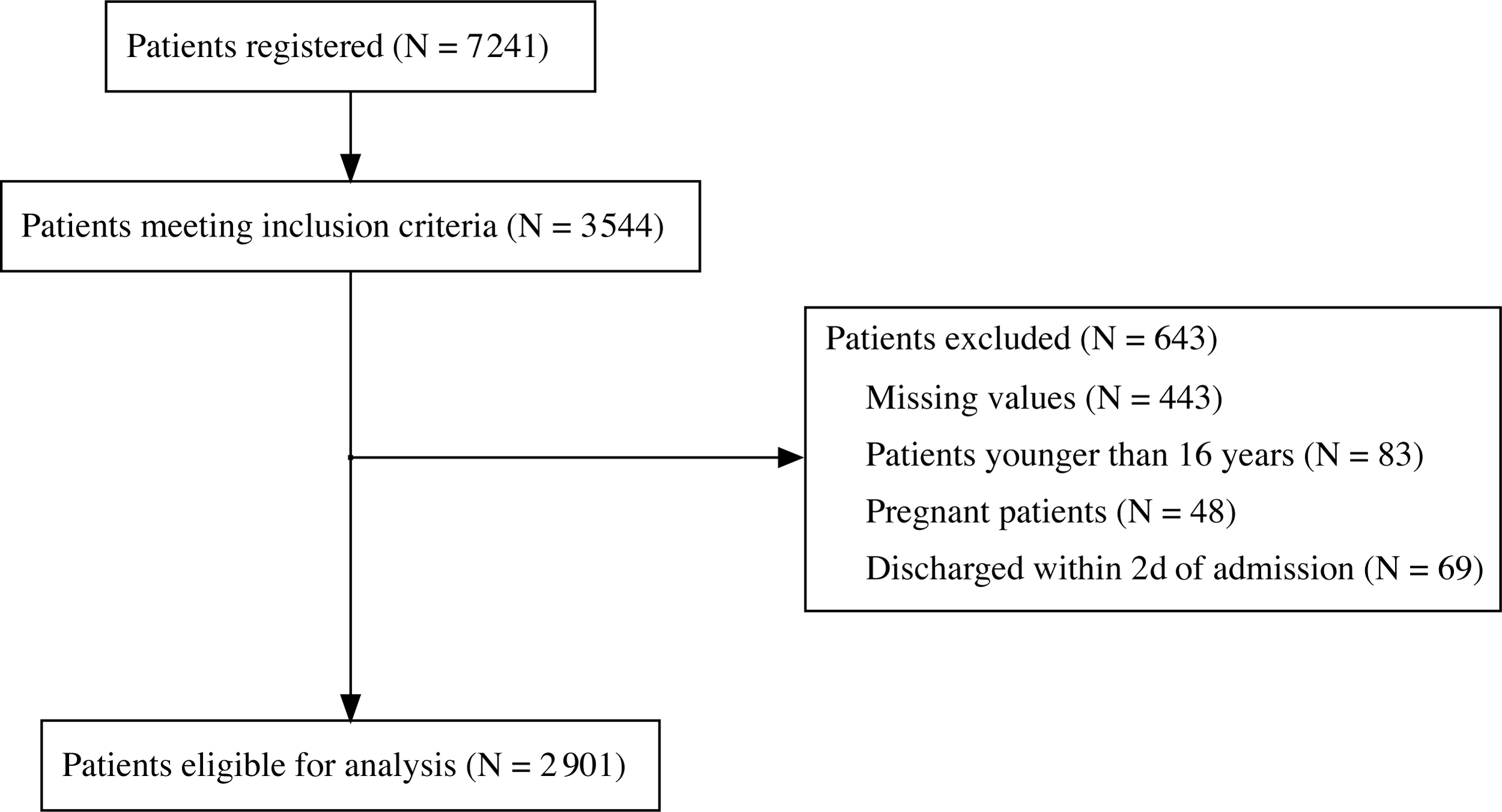
Flow diagram of the patient selection process. Abbreviations:

The GLMM established in this study showed satisfying prediction accuracy with an AUROC of 0.877±0.037 (Supplementary Figure 1). The results of the main analysis, in which the effect of endovascular treatment was estimated using this prediction model, are shown in Figure 2A. The adjusted odds ratio of in-hospital mortality was 1.45 [95% CI: 0.74–2.16, p = 0.21], showing no favorable outcome from this treatment. Regarding mRS, the adjusted odds ratio was 0.89 [95% CI: 0.56–1.23, p = 0.75], which cannot be construed as showing that endovascular treatments benefit patients with CVT.

**Figure 2:**
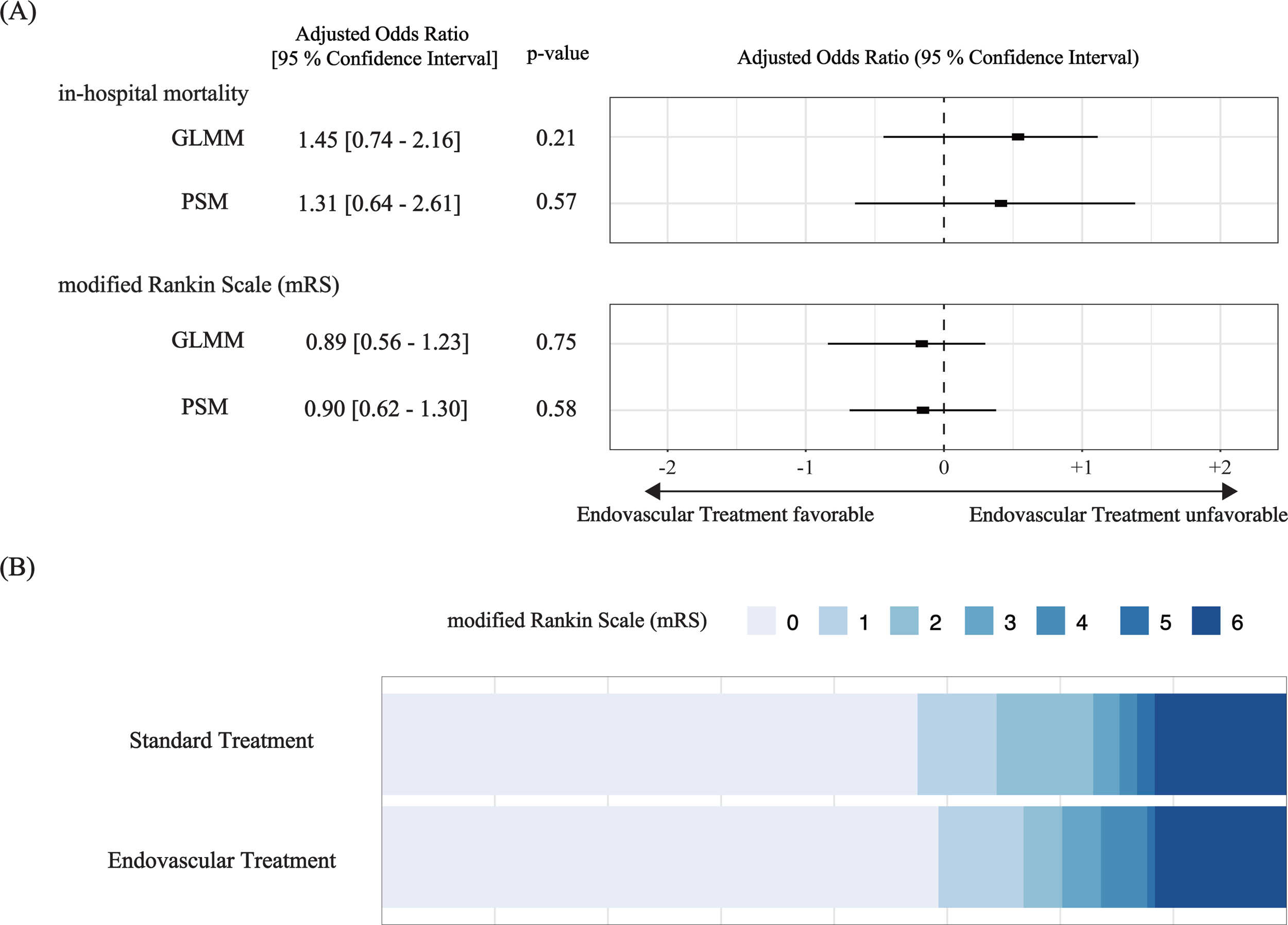
Outcome comparison between the endovascular and standard treatment groups. (A) Forest plot comparing the in-hospital mortality and modified Rankin Scale (mRS) of patients between the endovascular and standard treatment groups. (X-axis is presented in log scale). (B) mRS scores at the time of discharge. Abbreviations: GLMM, generalized linear mixed model; PSM, propensity score matching

The in-hospital mortality after the propensity score matching was 17/240 (7.1%) in the endovascular treatment group and 15/240 (6.2%) in the standard treatment group, consistent with the results estimated using the forementioned GLMM (adjusted odds ratio of 1.31 [95% CI: 0.64–2.61, p = 0.57] for in-hospital mortality and 0.90 [95% CI: 0.62–1.30, p = 1.30] for mRS), as shown in Figure 2A. The mRS scores of both groups at the time of discharge are shown in Figure 2B. The intercranial complication posthospitalization was 2/240 (0.8%) in the endovascular treatment group and 3/240 (1.2%) in the standard treatment group. No patients in either group had intracranial hemorrhage.

To identify the subpopulation that may benefit from endovascular treatment, we stratified the study cohort into three groups according to disease severity. As shown in Figure 3, a slight tendency toward favorable outcomes following endovascular treatments was observed in the low- and medium-risk groups, whereas unfavorable outcomes were reported in the high-risk group. However, neither of these results were conclusive (low-risk group: in-hospital mortality 0.36 [95% CI: 0.08–1.78, p = 0.27], mRS 0.31[95% CI: 0.09–1.09]; medium-risk group: in-hospital mortality 0.30 [95% CI: 0.04–2.36, p = 0.61], mRS 0.84 [95% CI: 0.27–2.60, p = 1.00]; high-risk group: in-hospital mortality 5.56 [95% CI: 0.81–37.19, p = 0.20], mRS 1.35 [95% CI: 0.57–3.19, p = 0.52]).

**Figure 3:**
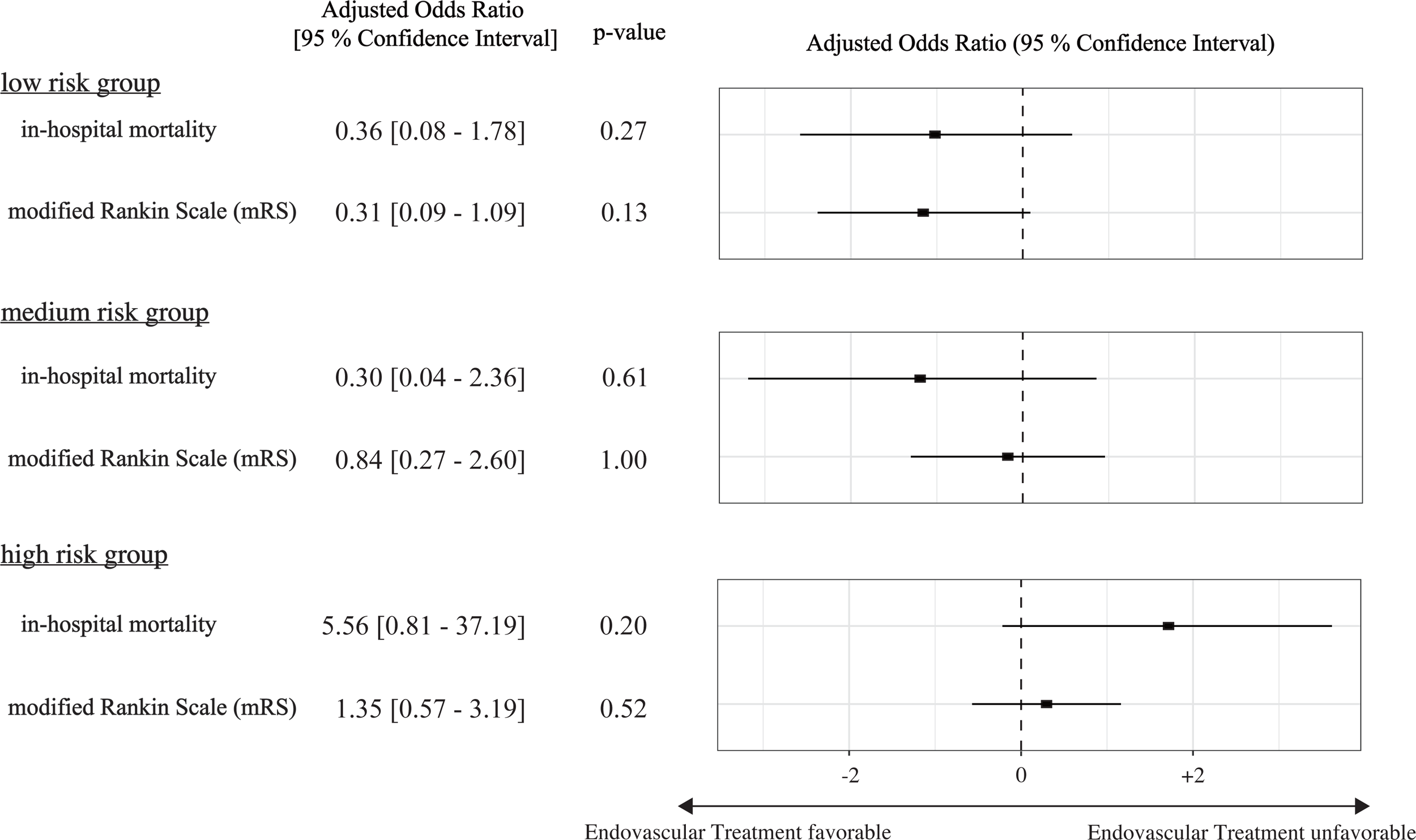
Outcome comparison among groups classified by disease severity. Case severities were stratified using the prediction model, while treatment effects were estimated using the propensity score matching. (X-axis is presented in log scale).

Figure 4 shows an additional analysis accounting for technological advancements, with subgroups stratified by the fiscal year of admission. The results of this subgroup analysis did not demonstrate any preference among endovascular treatment groups. Furthermore, as this study population was biased toward older patients, we also conducted an analysis using a limited population of patients aged <50 years; however, we found no evidence of favorable outcomes in the endovascular treatment group (in-hospital mortality 2.32 [95% CI: 0.73–19.30, p = 0.113], mRS 1.34 [95% CI: 0.57–1.79, p = 0.92]). These results are shown in Supplementary Figure 2.

**Figure 4:**
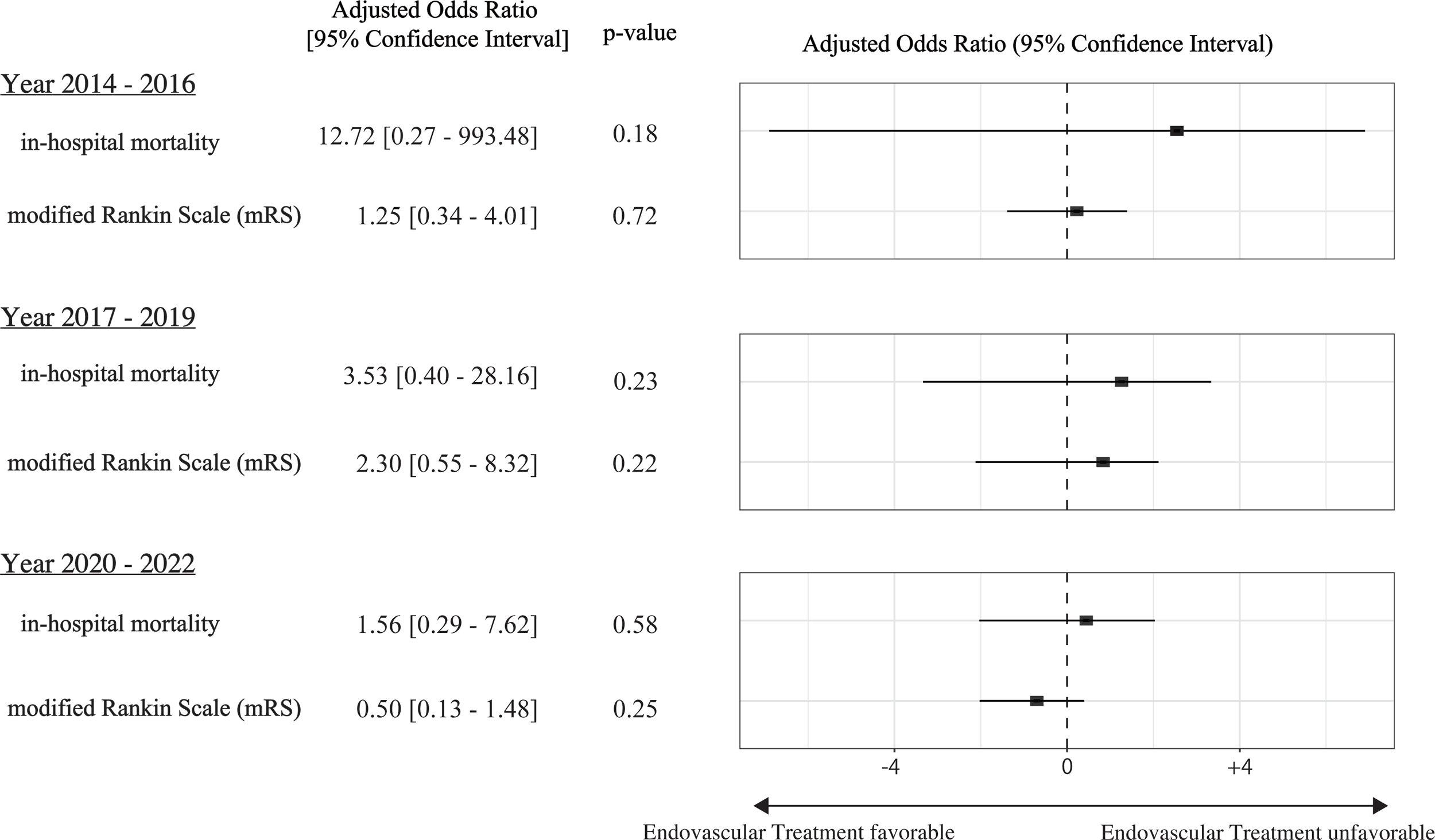
Outcome comparison among groups categorized by the year of admission. Treatment effects were estimated using a generalized linear mixed model. (X-axis is presented in log scale).

## Discussion

This study examined the efficacy of endovascular therapy in patients with CVT and found no benefits in terms of survival or neurological outcomes. We explored a subpopulation that might potentially benefit from this treatment; however, none of the results suggested favorability. Thus, our results, together with multiple estimation methods, did not suggest the efficacy of this treatment, ensuring robustness through multiple analyses.

Conflicting evidence exist regarding the use of endovascular treatments in patients with CVT. This was once welcomed with great enthusiasm and supported by positive evidence.^9,15^ However, a nationwide retrospective study analyzing 49 952 patients showed antagonistic results, indicating higher mortality in the endovascular treatment group.^16^ Furthermore, a multicenter randomized control trial was prematurely terminated due to futility.^10^ Nonetheless, these findings are insufficient for abandoning this treatment option. The prediction model of the aforementioned retrospective study was not sufficiently high (AUC = 0.75) and only a few variables were adjusted, raising the suspicion that the negative results may have been caused by unadjusted confounders. However, the randomized controlled trial was arranged from a sanguine perspective, aiming to detect no less than an absolute difference of 20%, resulting in enrolling only 34 patients in both arms. Considering the population size, a reliable subgroup analysis was impractical. The present study serves as a complement in this respect, as the study population included no less than 2901 patients, with patient severity being prudently adjusted, followed by a few subgroup analyses.

A recent systematic review, which included 405 patients from a randomized controlled trial and 20 observational studies, concluded that routine incorporation of endovascular therapy is not recommended,^17^ but with a reservation condition for severe patients. However, in the present study, favorable outcomes were observed neither in severe cases (Figure 3) nor in the most recently presented cases (Figure 4).

Of note, CVT has been previously reported to be much more common in women than in men^6,18^ and relatively rare in older patients.^19^ However, this tendency was not observed in the current study. We deduced this difference to several conditions unique to the Japanese population: (1) an aging population, (2) uncommon use of oral contraceptives,^20^ and (3) pregnant and puerperium patients being recorded differently in the database. To confirm the external validity of the present study while considering these reasons, we conducted a subgroup analysis limiting patients to those aged <50 years. Nonetheless, the obtained result was essentially equivalent to that of the main analysis (Figure 5).

Interpreting the results of the present study was arduous. Despite contradictory results regarding the benefits of endovascular treatments, no explicit complications were observed. As the safety of endovascular therapy has been repeatedly reported,^11,21^ treatment abandonment may be premature. Hence, more detailed research is required to reach definitive conclusions.

This study had some limitations. First, the diagnosis in this study was based on the ICD-10 code recorded in the database, which may be considered less definitive compared with the diagnosis in prospective investigations. However, a preceding study ensured the accuracy of the DPC database, showing that its specificity is above 96%.^22^ Second, the analysis was performed only for the patients who were hospitalized more than 2 d after admission. In total, 69 patients were discharged before this period, which is not likely to have affected the results of the study; however, the consequences of this bias have not been quantified. Third, the clinical efficiency of endovascular treatments in patients with CVT was measured, showing no observable improvement in terms of mortality and mRS scores. However, other measures, such as the occurrence of seizures and acute renal failure, were not evaluated. Fourth, the possibility of confounding factors remains, although the predictive model is well-validated. Nevertheless, this study provided a higher level of evidence than that provided in previous studies, as it was conducted in a large population and the analysis minimized the effect of confounding factors. In conclusion, no clinical benefit of endovascular treatments in patients with CVT was detected in the present study; however, further evidence is warranted.

## Data Availability

The data are not publicly available due to privacy concerns regarding the research participants.

## Non-standard Abbreviations and Acronyms

AUROC: Area under the receiver operating characteristic
CVT: Cerebral venous thrombosis
DPC: Diagnosis Procedure Combination
GLMM: Generalized linear mixed model
ICD-10: International Classification of Diseases, Tenth Edition
mRS: Modified Rankin Scale
PSM: Propensity score matching
SOFA: Sequential Organ Failure Assessment

## Acknowledgments

We thank Editage (www.editage.com) for providing excellent assistance with the English language editing.

## Authors’ contributions

AS and KF contributed to the acquisition of data, AS conceived of and designed this study. AS analyzed and interpreted the data. HS confirmed the analysis, KM, HS and KF drafted the manuscript. All of the authors reviewed and discussed the manuscript. KM and KF revised the manuscript for important intellectual content. All of the authors read and approved the final manuscript.

## Sources of Funding

None.

## Conflict of interest

None.

## Supplementary material

Supplementary Figure 1: Receiver operating characteristic curves of risk adjustment in the established model and validation cohort. The deviation of the model accuracy was estimated using the bootstrap method (N = 1000)

Abbreviations: AUROC, area under the receiver operating characteristics curve

Supplementary Figure 2: Outcome comparison in the subpopulation of patients aged <50 years.

Treatment effects were estimated using a generalized linear mixed model. (X-axis is presented in log scale).

## Notes

### Competing Interest Statement

The authors have declared no competing interest.

### Funding Statement

No funding was provided for this study.

### Author Declarations

Institutional Review Board of Tokyo Medical and Dental University

